# Promoting Sleep Duration in the Pediatric Setting Using a Mobile Health Platform: A Randomized Optimization Trial

**DOI:** 10.1101/2023.01.04.23284151

**Authors:** Jonathan A. Mitchell, Knashawn H. Morales, Ariel A. Williamson, Abigail Jawahar, Lionola Juste, Mary Ellen Vajravelu, Babette S. Zemel, David F. Dinges, Alexander G. Fiks

## Abstract

**Objective:** Determine the optimal combination of digital health intervention component settings that increase average sleep duration by ≥30 minutes per weeknight.

**Methods:** Optimization trial using a 2^5^ factorial design. The trial included 2 week run-in, 7 week intervention, and 2 week follow-up periods. Typically developing children aged 9-12y, with weeknight sleep duration <8.5 hours were enrolled (N=97). All received sleep monitoring and performance feedback. The five candidate intervention components (*with their settings to which participants were randomized*) were: 1) sleep goal (*guideline-based or personalized*); 2) screen time reduction messaging (*inactive or active*); 3) daily routine establishing messaging (*inactive or active*); 4) child-directed loss-framed financial incentive (*inactive or active*); and 5) caregiver-directed loss-framed financial incentive (*inactive or active*). The primary outcome was weeknight sleep duration (hours per night). The optimization criterion was: ≥30 minutes average increase in sleep duration on weeknights.

**Results:** Average baseline sleep duration was 7.7 hours per night. The highest ranked combination included the core intervention plus the following intervention components: sleep goal (either setting was effective), caregiver-directed loss-framed incentive, messaging to reduce screen time, and messaging to establish daily routines. This combination increased weeknight sleep duration by an average of 39.6 (95% CI: 36.0, 43.1) minutes during the intervention period and by 33.2 (95% CI: 28.9, 37.4) minutes during the follow-up period.

**Conclusions:** Optimal combinations of digital health intervention component settings were identified that effectively increased weeknight sleep duration. This could be a valuable remote patient monitoring approach to treat insufficient sleep in the pediatric setting.

## Introduction

Insufficient sleep duration impairs multiple dimensions of childhood development^1-4^, and is highly prevalent (53% of middle school and 65% of high school students)^5,6^. Prior research has shown that a 30 minute increase in sleep duration can lead to improvements in clinically relevant outcomes in children^7,8^. However, a meta-analysis reported that current sleep interventions for children increase average sleep duration by 10 minutes per night and most have been conducted in the school setting^9^.

It is recommended that pediatric clinicians screen for insufficient sleep and provide guidance at the point of care^10-13^; however, they lack effective tools and time at the point of care^8^. Mobile health platforms offer a solution to these barriers and theory-driven incentive strategies designed by behavioral economists can be integrated into mobile health platforms to enhance effectiveness. Of promise are loss-framed financial incentives that are motivational due to loss aversion^14^ and have been shown to enhance health behavior change in children^15^ and adults^15,16^. However, it is not known if behavioral sleep promotion and behavioral economic incentive components can be combined in a mobile health platform to promote sleep in children. To address this gap, we are using the Multiphase Optimization Strategy (MOST) framework to develop such a mobile health platform^17,18^.

The MOST framework includes three phases: *preparation, optimization*, and *evaluation*^17^. Prior to this study, we completed the *preparation phase*, demonstrating that it is feasible to deploy a mobile health platform with behavioral sleep promotion and behavioral economic incentive components for childhood sleep promotion^18^. In the present *optimization phase* study, we aimed to determine the optimal combination of candidate intervention components that increased weeknight sleep duration in children. We hypothesized that settings for the candidate intervention components would be identified that increased weeknight sleep duration by an average of ≥30 minutes.

## Methods

### Participants

Participants from southeastern PA and southern NJ were enrolled. Children aged 9-12 years were eligible; this avoided participants being exposed to earlier high school start times, and more advanced pubertal stages when sleep is impacted by the circadian process^19^. Participants had to have insufficient sleep, which was assessed by caregiver report (6.0 to 8.5 hours asleep on school nights on a screening questionnaire) and sleep tracker (<8.5 hours asleep on school nights during the run-in phase). Participants had to have access to a tablet or smartphone so they could transmit their sleep data. Only a single child per family could participate and they had to sleep in their own bed. We enrolled typically developing children and excluded those with a known sleep disorder (e.g., sleep apnea), psychiatric disorder diagnosis (e.g., ADHD, depression, anxiety, an eating disorder), a musculoskeletal or neurological disorder that limits physical movement, or any medication use known to affect body weight and/or sleep. The Children’s Hospital of Philadelphia’s (CHOP) Institutional Review Board approved this study, and it is registered at clinicaltrials.gov (NCT03870282).

### Timing of Data Collection

Data were collected between March 2019 and December 2020. In March 2020, PA and NJ issued curtailments to help prevent coronavirus disease 2019 transmission, including school closures. By fall 2020, some curtailments were eased, and some districts allowed hybrid schooling. We categorized participants based on when they completed the study: 1) Spring-Fall 2019 semesters, 2) Spring 2020 semester, or 3) Fall 2020 semester.

### Mobile Platform

Way to Health is an automated information technology platform that integrates wireless devices, randomization, digital messaging, and secure data capture^20,21^. A Fitbit sleep tracker (Flex 2 or Inspire) was used to measure sleep duration in the home setting; this is a single sensor device with a proprietary algorithm used to estimate sleep from locomotor data collected by the accelerometer. The sleep data were transferred to the Way to Health platform using an application programming interface. Fitbit devices have been validated against polysomnography in children and the validity metrics are comparable to traditional actigraphy devices (i.e., high sensitivity and moderate specificity)^22-24^.

### Study Design

All participants completed a 2-week run-in period. Baseline sleep duration was calculated using data from the second run-in week. If two or more nights of data were provided and average sleep duration was <8.5 hours on weeknights (Sun-Thurs) participants were randomized to a study condition for a 7-week intervention period, followed by a 2-week follow-up period. Participants were randomized using block randomization and a random number generator in the Way to Health platform. Blinding was not used.

All received the core intervention: sleep tracking and weekly performance feedback. Weekly performance feedback text messages were sent each Sunday during the intervention period and included tailored supportive feedback to maintain strong performances or to improve upon weaker performances. By default, the messages were sent to caregivers. A 2^5^ factorial design was used to test the effectiveness of five candidate intervention components (Supplementary Figure 1): 1) sleep goal (*guideline-based or personalized*); 2) child-directed loss-framed financial incentive (*inactive or active*); 3) caregiver-directed loss-framed financial incentive (*inactive or active*); 4) screen time reduction messaging (*inactive or active*); and 5) daily routine messaging (*inactive or active*). The conceptual model is provided in Supplementary Figure 2.

The sleep goal component provided a guideline-based (≥9.25 hours) or personalized (≥45 minutes greater than baseline) goal. Middle school aged children are recommended to sleep for 9-12 hours each day^25^, justifying a guideline-based goal. However, Self-Efficacy Theory predicts that children will be more motivated if they believe that they can attain a given goal^26^. A personalized goal approach may minimize any differential self-efficacy arising from varied baseline sleep duration.

Loss-framed incentives can help promote behavior change^15,16,18^. Based on Prospect Theory, loss-framed incentives are motivational because of loss aversion^14^. However, in the pediatric context we do not know if it is best to direct loss-framed incentives to caregivers, children or both^27^. We therefore included child-directed and caregiver-directed loss-framed incentives as candidate components. If activated, the incentive components provided participants with a $70 endowment, with $2 deducted if the sleep goal was not achieved on weeknights during the intervention period. Money was dispensed in weekly installments (i.e., up to $10 per week). The $2 value matches the value used in a prior health behavior study^15^.

Reducing screen time is considered important for helping youth extend their sleep^28-31^. If activated, the screen time reduction messaging component provided families with key information on why electronic screens are detrimental to sleep; provided solutions to help reduce screen time; and provided supportive messages to encourage electronic device use reduction. Twelve messages were sent to caregivers (4 per week during intervention weeks 1, 3 and 5).

Observational studies consistently show that children are more likely to sleep sufficiently if their caregivers set consistent bedtimes^32-36^, and if they manage time spent doing extracurricular activities^37,38^. If activated, the messaging component to establish daily routines provided families with key information on why routines are important for optimal sleep and strategies on how to develop bedtime and daytime routines. Twelve messages were sent to caregivers (4 per week during intervention weeks 2, 4 and 6).

### Optimization Criterion

A 30-minute increase in sleep duration improves clinical outcomes in youth^7,8^. We considered the candidate component settings to be optimal if they helped to increase average weeknight sleep duration by ≥30 minutes.

### Survey Data

Participants completed questionnaires and the following items are included for descriptive purposes: age, race/ethnicity, and sex (male or female).

### Statistical Analysis and Power

The primary analyses proceeded as intent-to-treat. Mixed effect linear models, with random intercepts and slopes, and an unstructured covariance structure, were used to model changes in weeknight sleep duration. To assess for individual component contributions (i.e., main effects), candidate component by week interactions were included as fixed effects. To assess if combinations of component settings contributed to changes in weeknight sleep duration (i.e., interaction analysis), we assessed the full model that included all candidate component main effects and interactions as fixed effects. We then used the following model reduction process: 1) kept all interactions with *P*-values ≤0.15 for ≥4 intervention weeks; 2) refit the model; and 3) ranked the remaining interactions based on the predicted changes in sleep duration during the intervention period. These analyses were performed using Stata version 17.0 (StataCorp, College Station, TX). We used the MOST R package to determine that we needed a minimum sample size of 65 participants to detect a 30-minute difference in sleep duration at alpha=0.05 and power=0.8. To help ensure a balanced design, and to account for participant dropout, we aimed to enroll and randomize 96 participants to achieve 48 participants per component level.

## Results

Ninety-seven participants were randomized with equal distribution across component levels (Figure 1). The average age was 11.5y, and the sample was 51% female, 29% Black, and 56% White (Table 1). There were no differences in sociodemographics, collection period, or sleep duration by the component levels at baseline (Table 1). Nights of sleep data captured were similar across sociodemographic factors, collection period, and component levels, except for 1-3 fewer nights per week of data captured for Black compared to White participants (Figure 2). Average weeknight sleep duration was 7.7 hours per night at baseline (Table 1). Overall, weeknight sleep duration increased from baseline by an average of 24 (95% CI: 17, 32) and 21 (95% CI: 11, 32) during the intervention and follow-up periods (Supplementary Figure 3).

**Table 1.**
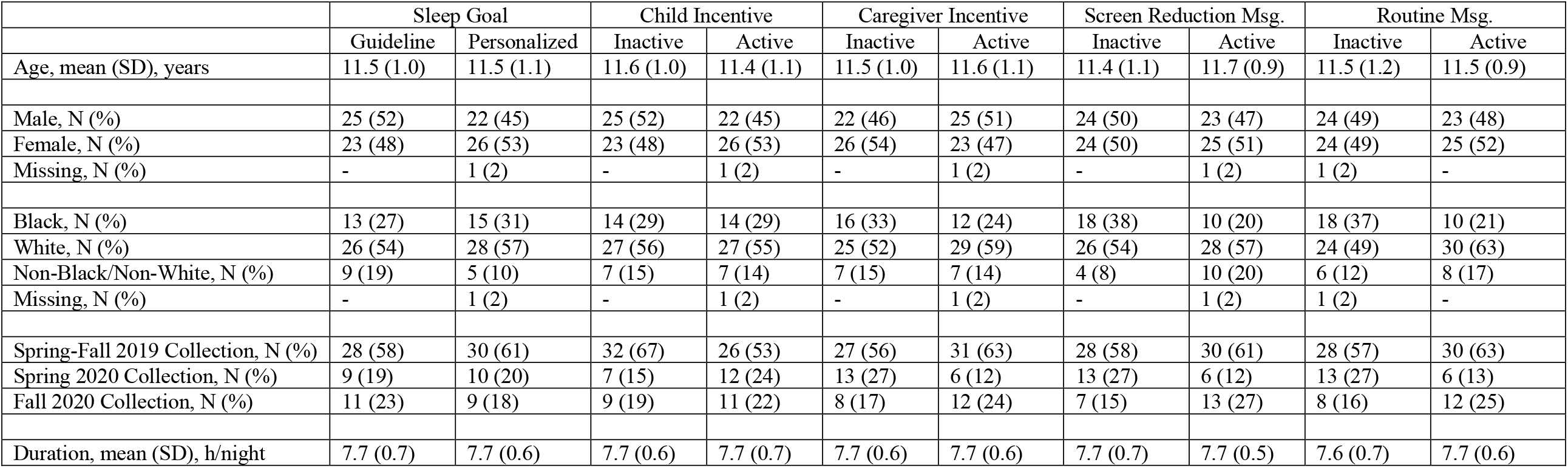
Baseline characteristics by candidate component settings

**Figure 1.**
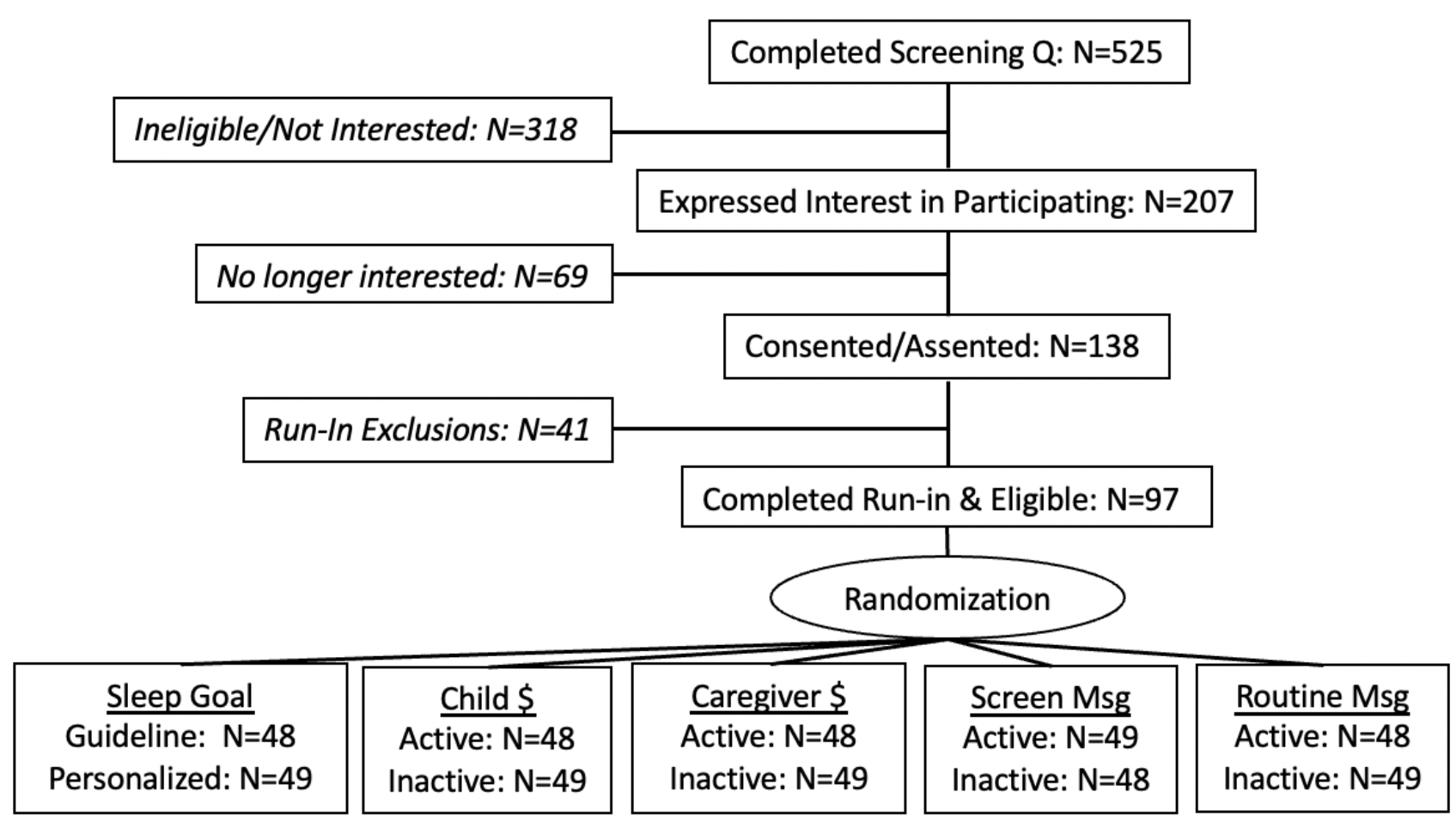
Participant flow diagram. Abbreviations: Q, questionnaire. All 97 participants randomized were included in the analyses.

**Figure 2.**
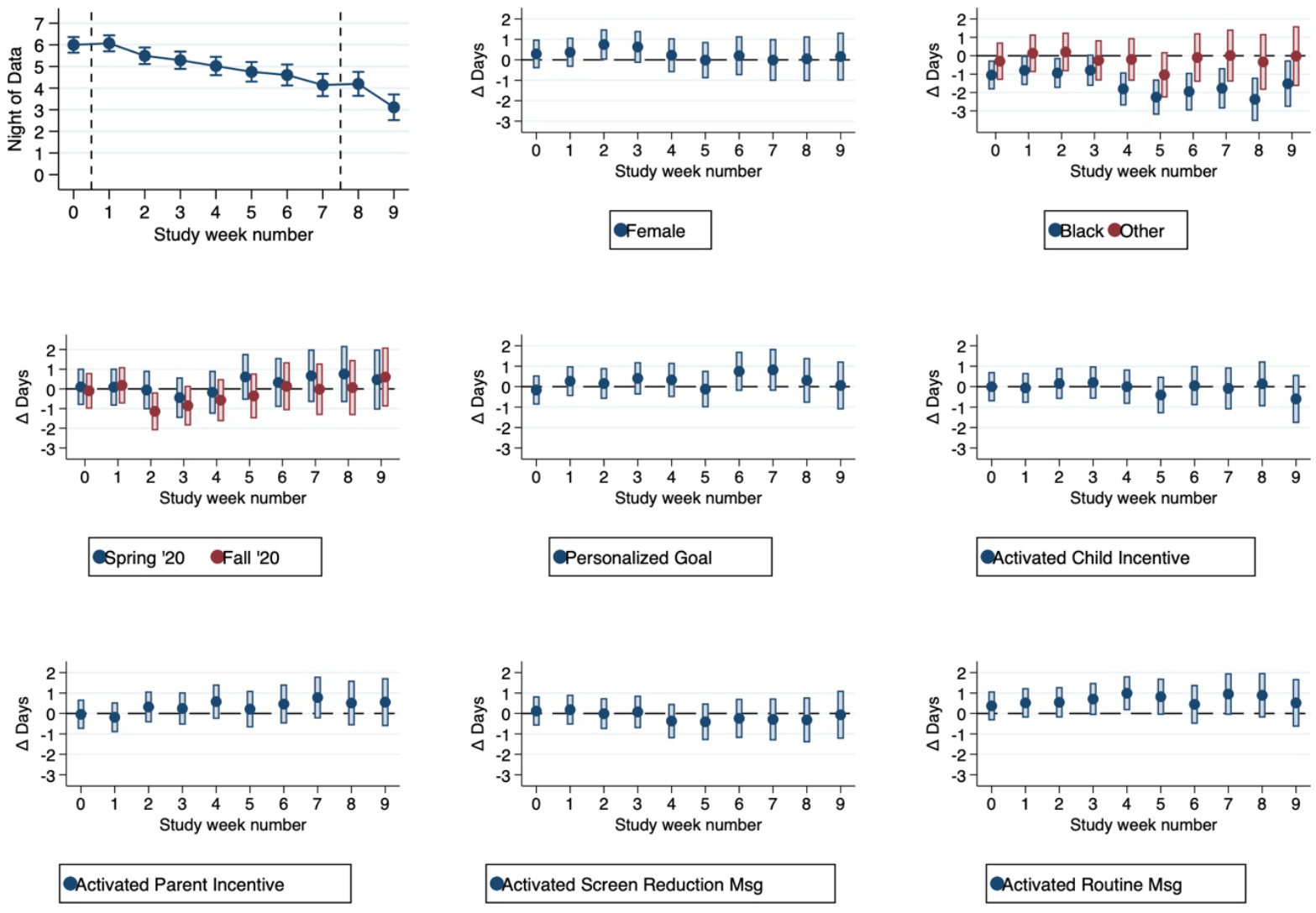
Average nights of data captured by study week for sociodemographic factors, data collection period, and candidate component levels. The horizontal lines at 0 indicates no difference between nights of data collected by sociodemographic factors, data collection period, and candidate component levels.

We did not detect study week by component interactions, meaning changes in sleep duration post baseline were comparable for either component setting for a given candidate component, with only modest favorable effects detected (Figure 3). Specifically, changes in weeknight sleep duration from baseline were: 1) more favorable for the guideline sleep goal setting during the intervention (+11 minutes, 95% CI: −4, 26) and follow-up periods (+6 minutes, 95% CI: −14, 26); 2) less favorable when the child-directed loss-framed incentive was activated during the intervention period (−5 minutes, 95% CI: −19, 10) but more favorable during the follow-up period (18 minutes, 95% CI: −3, 38); 3) more favorable when the caregiver-directed loss-framed incentive was activated during the intervention during the intervention period (+7 minutes, 95% CI: −8, 22) but less favorable during the follow-up period (−19 minutes, 95% CI: −40, 1); 4) less favorable when screen time reduction messaging was activated during the intervention (−4 minutes, 95% CI: −19, 11) and follow-up periods (−6 minutes, 95% CI: −27, 14); and 5) less favorable when routine messaging was activated during the intervention (−5 minutes, 95% CI: − 20, 10) and follow-up periods (−5 minutes, 95% CI: −25, 16). No setting for a single candidate intervention component achieved the optimization criterion.

**Figure 3.**
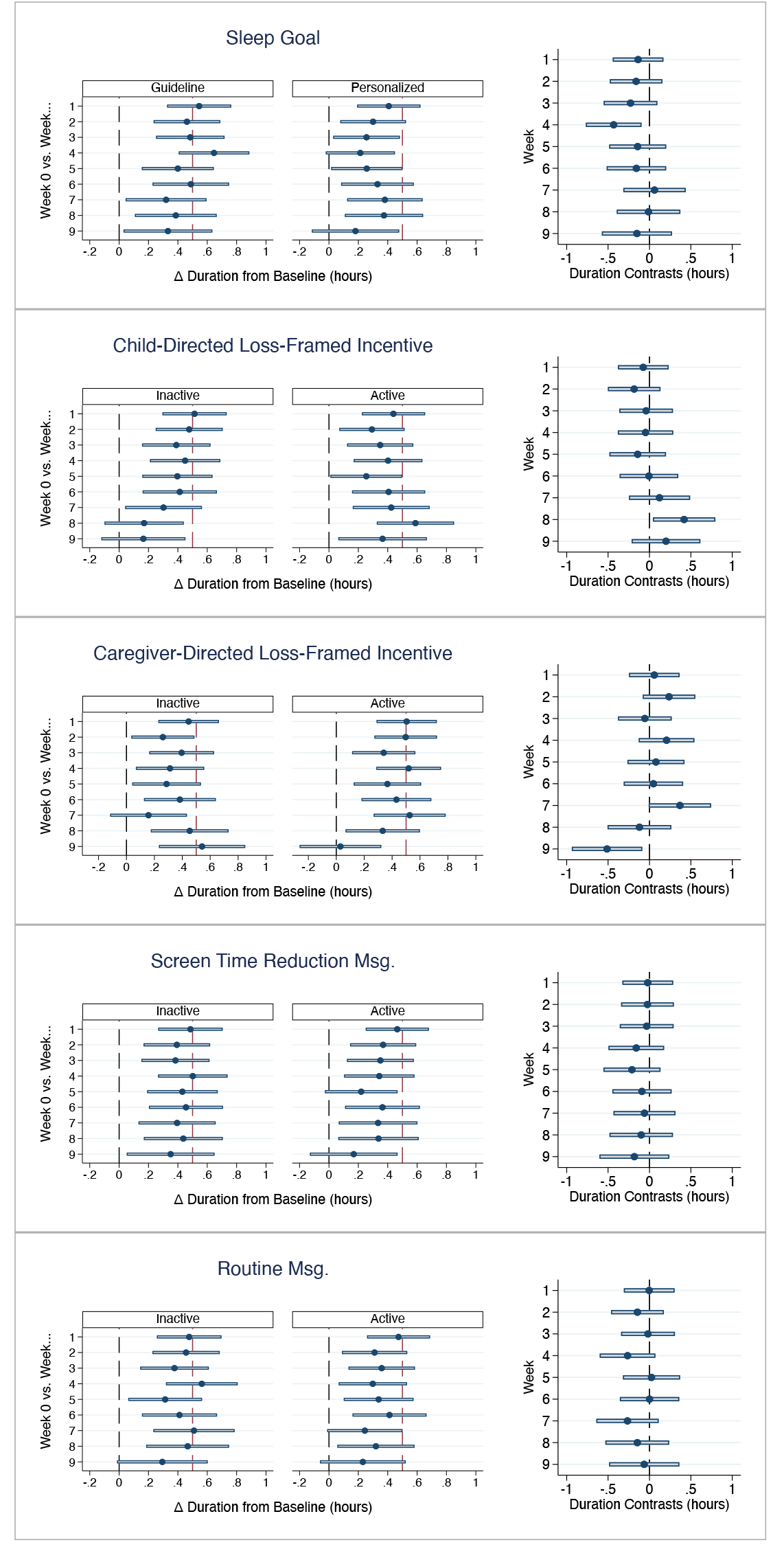
Candidate component main effects. The left column shows the average change in sleep duration from baseline by subsequent study weeks by each setting. The right column shows the difference in sleep duration change from baseline between the component settings by study week.

For the component interaction analysis, the model reduction process yielded five component combinations, of which two achieved the optimization criterion (Table 2). The highest ranked combination included the core intervention plus the following components: caregiver-directed loss-framed incentive, messaging to reduce screen time reduction, and messaging to establish daily routines; either sleep goal setting was equally effective. Weeknight sleep duration increased by an average of 39.6 minutes (95% CI: 36.0, 43.1) and 33.2 minutes (95% CI: 28.9, 37.4) during the intervention and follow-up periods, corresponding to a 20% increase in the number of weeknights with sufficient sleep duration (Table 2). The increase in sleep duration from baseline occurred in week 1 and was maintained throughout the intervention period (Figure 4, panels A and B).

**Table 2.** Component combinations and ranked changes in weeknight sleep duration

| | | | | | | Change in Weeknight Sleep Duration (minutes) | | | | Change in Weeknight Sleep Duration $\geq 9$h (proportion) | |
| --- | --- | --- | --- | --- | --- | --- | --- | --- | --- | --- | --- |
|  | Core Intervention + Component Combinations... |  |  |  |  | Intervention Period |  | Follow-up Period |  | Intervention Period | Follow-up Period |
| Rank | Goal | Caregiver \$ | Child\$ | Screen Msg | Routine Msg | Estimate | 95% CI | Estimate | 95% CI | Estimate | Estimate |
| 1 | G or P | Active | Inactive | Active | Active | 39.6 | 36.0, 43.1 | 33.2 | 28.9, 37.4 | 0.20 | 0.20 |
| 2 | G or P | Active | Inactive | Inactive | Active | 32.0 | 28.4, 35.6 | 26.6 | 22.4, 30.8 | 0.19 | 0.19 |
| 3 | G | Active | Inactive | Active | Inactive | 29.7 | 26.6, 32.8 | 23.8 | 20.1, 27.6 | - | - |
| 4 | G | Active | Active | Active | Inactive | 28.4 | 24.4, 32.5 | 19.1 | 14.2, 23.9 | - | - |
| 5 | G | Inactive | Active | Active | Inactive | 16.2 | 12.5, 19.9 | 10.6 | 6.2, 14.9 | - | - |

Abbreviations: G, guideline-based sleep goal; P, personalized sleep goal. Estimate (95% CI): average change in weeknight sleep duration in minutes from baseline. Model predicted average change in the proportion of weeknights with sleep duration $\geq 9$ hours from baseline was not calculated for combinations that failed to surpass the optimization criterion.

**Figure 4.**
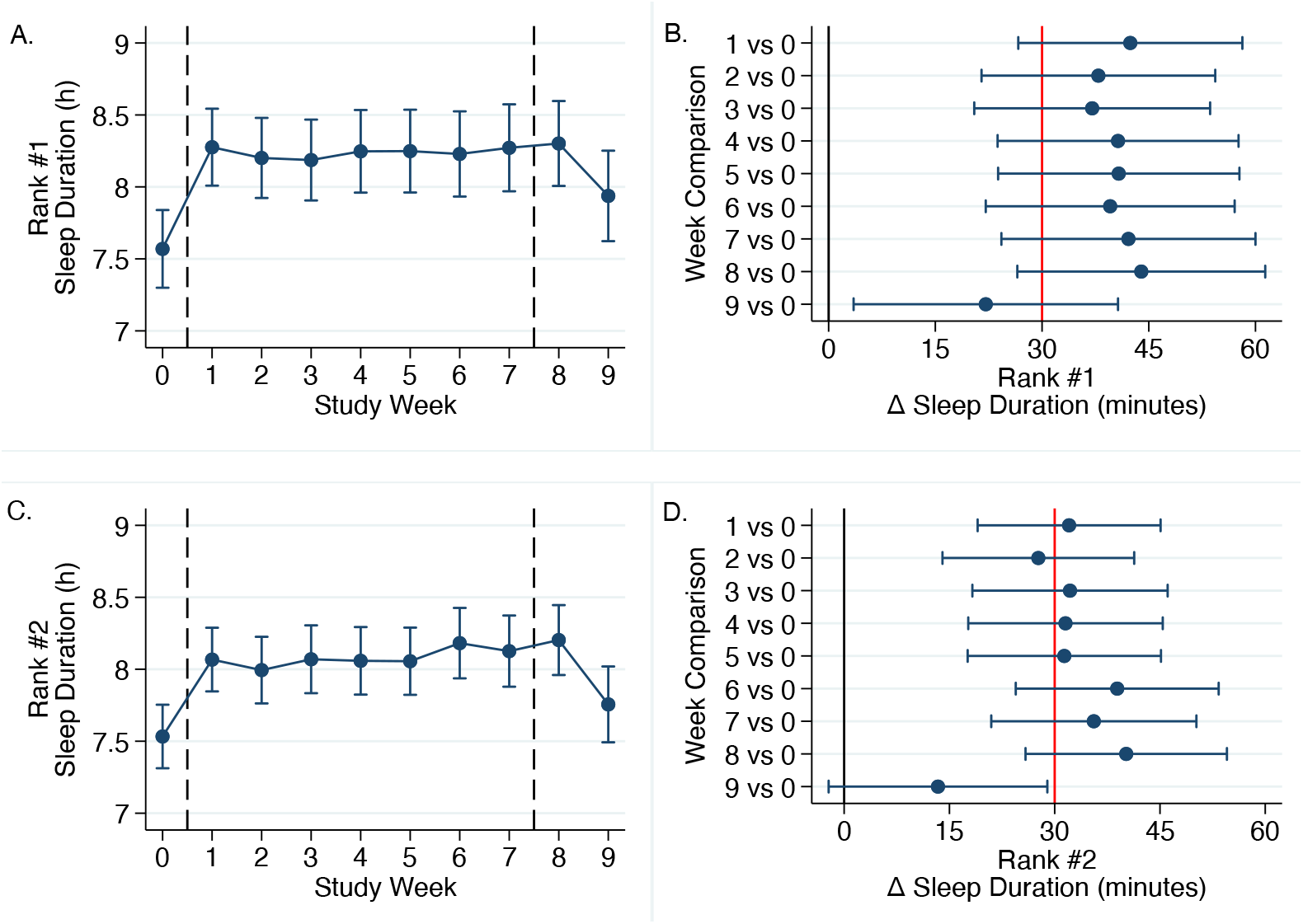
Predicted sleep duration by study week and predicted changes in weeknight sleep duration from baseline by ranked component combinations (panels A and B for rank #1 and panels C and D for rank #2). The dashed reference lines in panels A and C distinguish baseline, intervention, and follow-up periods. In panels B and D, the black reference lines distinguish changes in sleep duration from baseline above zero, whereas the red reference lines indicate the optimization criterion.

The second ranked combination included the core intervention plus the following components: caregiver-directed loss-framed incentive and messaging to establish daily routines; either sleep goal setting was equally effective. Sleep duration increased by an average of 32.0 minutes (95% CI: 28.4, 35.6) and 26.6 minutes (95% CI: 22.4, 30.8) during the intervention and follow-up periods, corresponding to a 19% increase in the number of weeknights with sufficient sleep duration (Table 2). Again, the increase in sleep duration from baseline occurred in week 1 and was maintained throughout the intervention period (Figure 4, panels C and D).

Component combinations ranked third and fourth were in proximity to the optimization criterion (Table 2). Whereas the fifth ranked combination was not in proximity to the optimization criterion (Table 2). The third, fourth, and fifth ranked combinations had common settings with respect to the core intervention, guideline-based sleep goal, and messaging to reduce screen time (Table 2). The activation and inactivation of the caregiver-and child-directed incentives distinguished these three combinations, revealing that the caregiver-directed loss-framed incentive alone (third ranked) was more effective than the child-directed loss-framed incentive alone (fifth ranked), and that having both loss-framed incentives activated did not enhance effectiveness (fourth ranked) (Table 2).

## Discussion

Pediatric care teams lack effective and efficient interventions to treat childhood insufficient sleep duration^10-12^. To address this gap, we are using the MOST framework to engineer a mobile health sleep promotion platform with behavioral sleep promotion and behavioral economic incentive components for the pediatric setting^18^. To the best of our knowledge, we are among the first to apply the MOST framework to develop a childhood sleep promotion intervention. In the present study, under the *optimization phase* of the MOST framework, we enrolled children with insufficient sleep duration into an optimization trial and found two combinations of component settings for our mobile health platform that increased weeknight sleep duration by ≥30 minutes. Our next step is to complete the *evaluation phase* of the MOST framework and determine if the optimized intervention package can effectively increase sleep duration using a randomized controlled trial design. While this process remains in development, this mobile health approach has the potential to be incorporated into the pediatric primary care setting and could prove to be a valuable tool as part of a sleep health workflow to help address insufficient sleep^39^.

There is evidence that mobile health approaches can be used to set and monitor the achievement of behavioral goals^40^, and that loss-framed incentives linked to goals enhance effectiveness, due to loss aversion^41-43^. Our previous *preparation phase* research supported this concept for sleep promotion in children when using a guideline-based sleep goal and a caregiver-directed loss-framed incentive^18^. In the present optimization trial, we found that both settings for the sleep goal component were comparable, but with the guideline setting being more favorable. The component interaction analysis revealed that both sleep goal settings were equally effective in the top two ranked combinations that achieved the optimization criterion, but the guideline-based sleep goal setting was more effective than the personalized sleep goal setting in the third and fourth ranked combinations that were in proximity to the optimization criterion. It would therefore be reasonable to adopt a guideline-based sleep goal as the default setting, with the option to switch to a personalized setting if desired.

Regarding the loss-framed incentives, we found the child-directed loss-framed incentive was not favorable during the intervention period and the interaction analysis confirmed that the child-directed loss-framed incentive component was redundant and would add an unnecessary cost. There is evidence that a loss-framed incentive directed at adolescents with type 1 diabetes improved blood glucose monitoring^15^, and a loss-framed incentive improved increased fruit consumption in children^44^. However, in the latter study the gain-framed incentive of equal value was equally effective^44^. Future research could investigate whether gain-or loss-framed incentives are more efficacious at promoting sleep in children, and further consideration could be given to the timing and magnitude of the incentive used, and if the incentive approach used to developmentally appropriate across childhood^45^.

Our main effect analysis revealed that when activated the caregiver-directed loss-framed incentive was more favorable during the intervention period (but not during the follow-up phase). Further, the interaction analysis revealed that the caregiver-directed loss-framed incentive was effective in the four top-ranked combinations of components. These data indicate that a caregiver-directed loss-framed incentive should be included in the mobile health platform. However, it is worth noting that longer-term follow-up is needed to assess for lasting effects especially since the caregiver-directed incentive is a source of extrinsic motivation and habit formation was not confirmed in our study. With respect to future implementation, several states have received waivers to enact Healthy Behavior Incentive Programs^46^, and private insurers are increasingly offering enrollees discounted premiums if behavioral targets, measured by wearable devices, are achieved^47^. These data indicate that a caregiver-directed loss-framed incentives for adhering a sleep goal has the potential to be implemented into pediatric healthcare and warrants specific study.

Our optimization trial included messaging components for establishing daily routines and reducing screen time. The former provided families with information and guidance on how to set bedtimes and establish pre-bedtime routines, as well as encouraging the adoption of daytime strategies that should be conducive to sleep promotion^38^. The latter provided information and guidance on managing screen time, especially use of electronic devices in the bedroom^48^. Both messaging components contributed to the top ranked combination that achieved the optimization criterion, and the messaging component encouraging daily routines also contributed to second ranked combination that achieved the optimization criterion. Our data therefore support including both messaging components, although the messaging component for reducing screen time could be removed without severely compromising effectiveness. This might be advantageous from a personalized messaging approach, which was identified as being desirable from our prior qualitative research^18^.

Our study has limitations. This was a single site study in one geographic location with a sample that was predominantly non-Hispanic White. We captured 1-3 days less per week on average among Black participants, and we were not able to test if the component combinations were equally effective across the sociodemographic spectrum, which has implications for promoting sleep health equity^49,50^. Our follow-up period lasted for two weeks, and a future study should determine the long-term impact of this mobile health approach on childhood sleep promotion. The commercial sleep tracker used had a single sensor (accelerometer) and a proprietary algorithm; a multi-sensor sleep tracker (e.g., accelerometer and heart-rate monitor) may improve the estimation of sleep and open-source algorithms would provide transparency for replication efforts.

In conclusion, optimal settings for the behavioral sleep promotion and behavioral economic components of our mobile platform were identified that increased sleep duration by ≥30 minutes. Specifically, we discovered that sleep tracking, performance feedback, a sleep goal, a caregiver-directed loss-framed incentive, and messaging to reduce screen time and establish daily routines are optimal settings for our mobile health approach that has potential to be implemented in the pediatric primary care setting.

## Data Availability

All data produced in the present study are available upon reasonable request to the authors

---

**Supplementary Figure 1.**
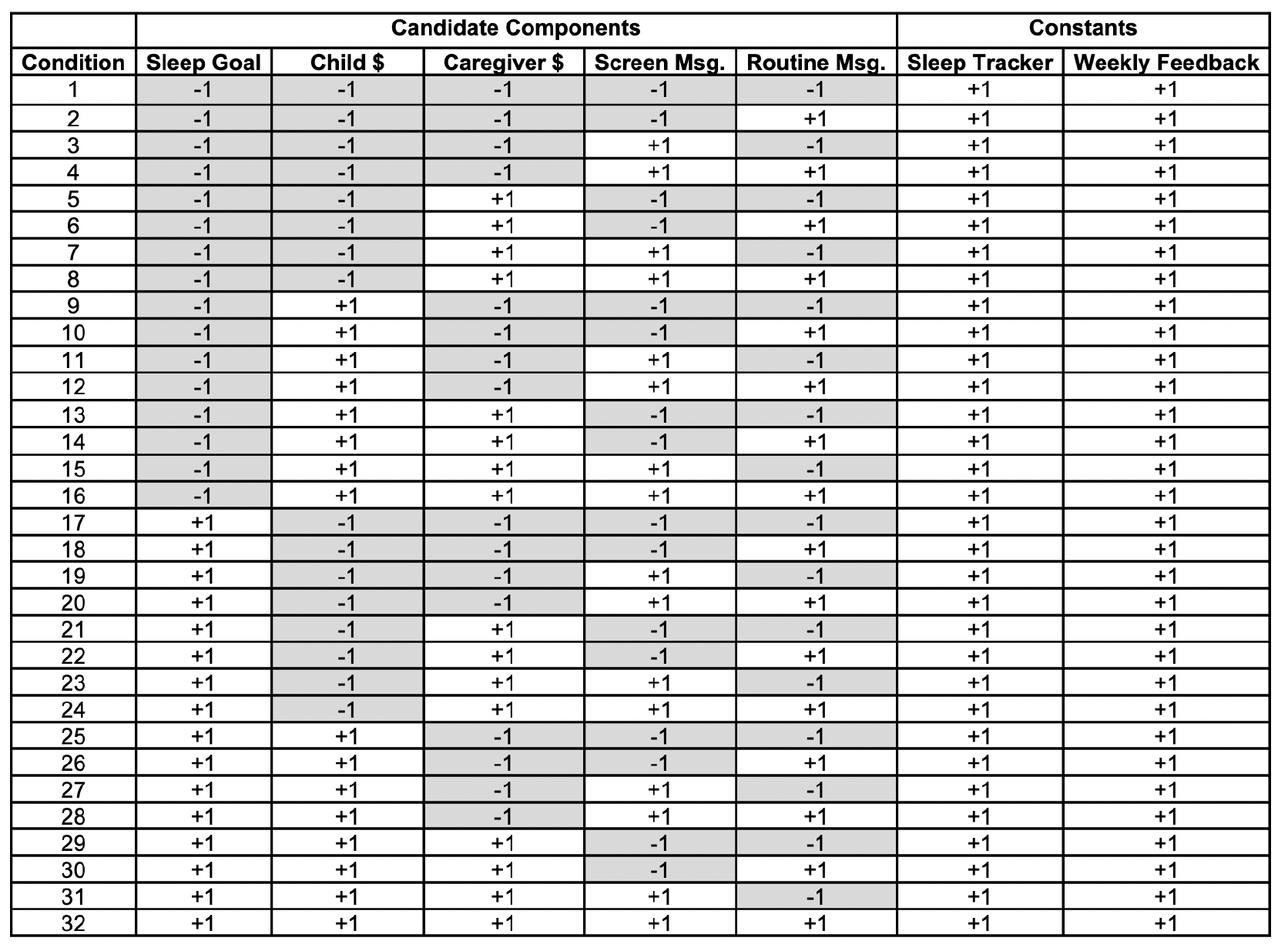
Factorial study design used in the optimization trial. Effect coding was used to distinguish the two levels of each candidate components (−1 for the guideline-based sleep goal and inactived settings, and +1 for the personalized sleep goal and activated settings). All participants recvied the constants (sleep tracking and weekly performance feedback).

**Supplementary Figure 2.**
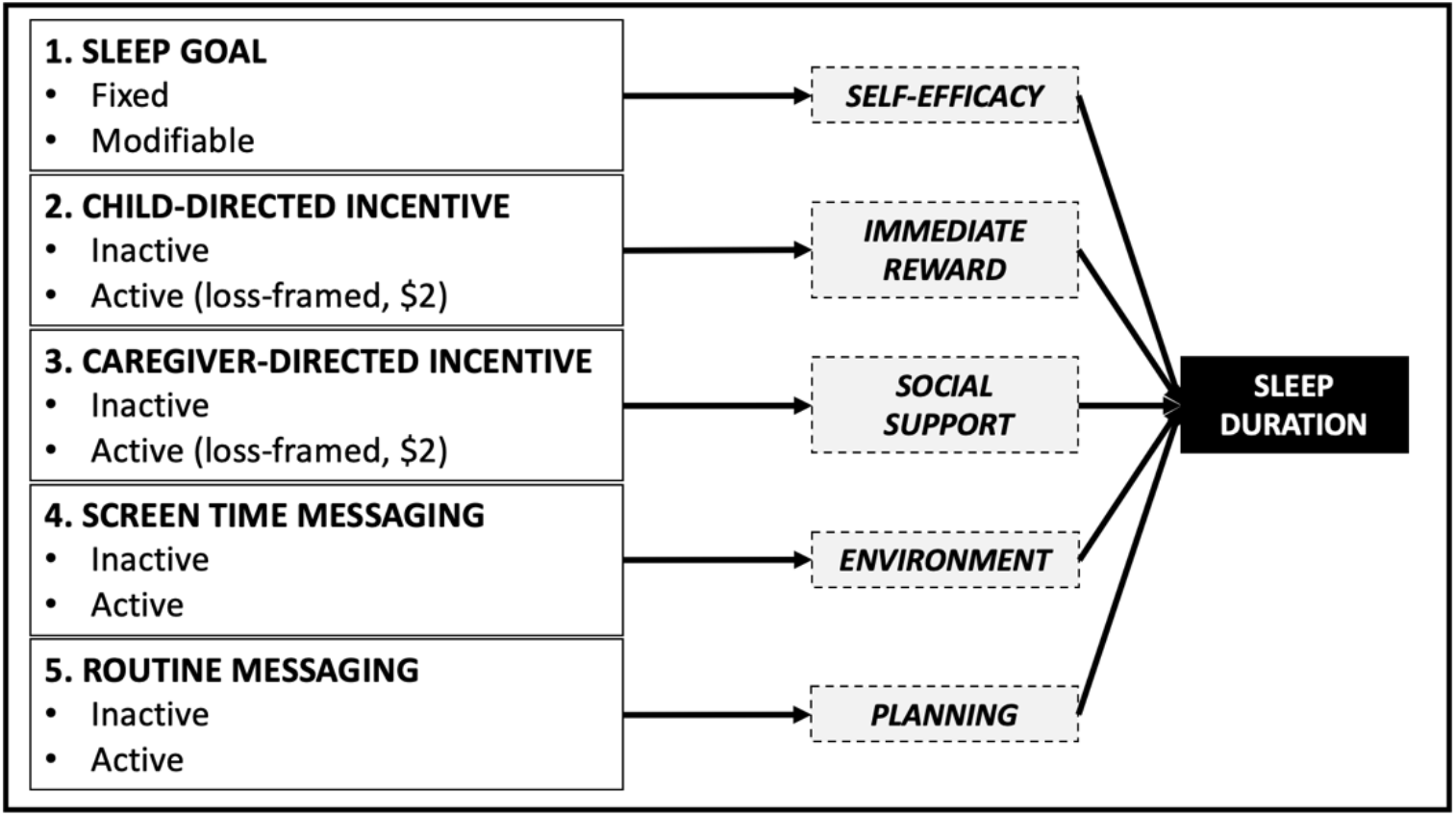
Conceptual model for the intervention components

**Supplementary Figure 3.**
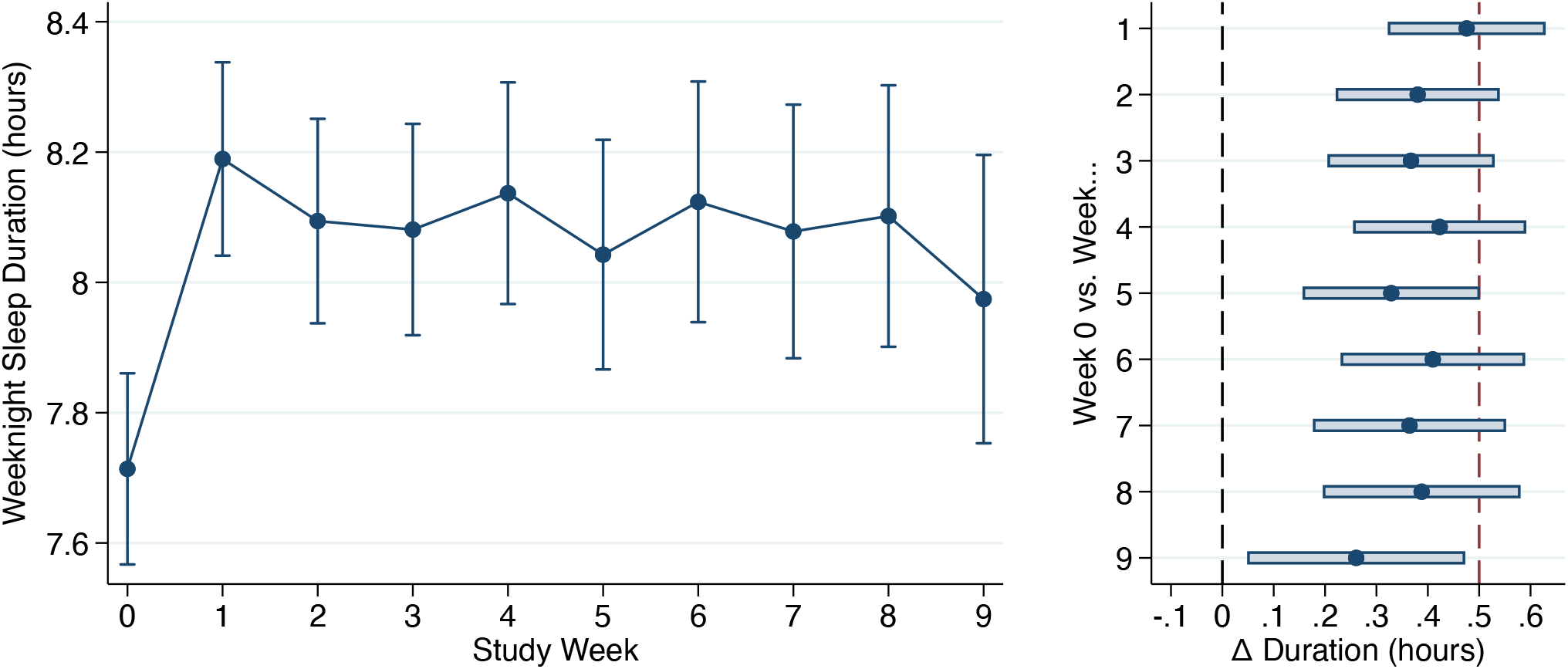
Overall change in sleep duration by study week in the entire sample.

